# Feasibility of penile erection detection through measurements of light reflection of hemoglobin of the penis

**DOI:** 10.1101/2024.05.06.24306762

**Authors:** Evelien Trip, Hille Torenvlied, Henk Elzevier, Rob Pelger, Jack Beck

## Abstract

Timely and accurate diagnosis of erectile dysfunction (ED) is crucial for effective treatment and early detection of health risks. There is a need for a non-invasive sensor for erection detection. This study aims to investigate the feasibility of penile erection detection through measurements of light reflection of hemoglobin (LRH) of the penis. A penile LRH sensor was developed to measure changes in hemoglobin levels of the flaccid and erected penis as a precursor to the development of a SpO2-sensor. Outcomes from penile duplex and LRH measurements in ED patients were compared in the flaccid state as well as during an erection caused by intravenous injection of alprostadil. Results of ten patients getting a full erection are considered. The results show a difference in LRH values during erections measured on the penis compared to a flaccid penis. This is the first human pilot study to demonstrate a higher LRH during the onset of an artificially induced erection in patients with ED, this tendency should be analyzed in larger studies. This is a promising advancement towards the further development of a much-needed non-invasive sensor that has the potential to revolutionize the diagnosis and management of ED.

## 1. Introduction

Erectile dysfunction (ED) is defined as the persistent or recurrent inability to attain or maintain an erection sufficient to perform sexual activity [1]. The global prevalence of ED varies between 3% and 77% and is associated with higher age [2]. The etiology of Erectile Dysfunction (ED) encompasses both psychological and organic factors. Psychological issues can often contribute significantly to the manifestation of ED. Organic ED can broadly be categorized into vascular, neurogenic, iatrogenic, or endocrine causes, or a combination [3]. Understanding these diverse factors is crucial in comprehensively assessing and addressing the complexities of this condition. ED influences physical and psychosocial well-being [4,5]. ED has also shown to be an important sentinel marker of cardiovascular conditions [3,6,7]. Thus, timely and accurate diagnosis of ED is essential for both adequate treatment of the dysfunction as well as early identification of other potentially life-threatening conditions and possibly the prevention of cardiovascular disease [7-10]. Given that vasculogenic erectile dysfunction often precedes coronary artery disease, myocardial infarction and strokes [9].

Normal erectile functioning depends on three important factors: hemodynamic interaction of cavernosal arterial inflow, perfusion pressure and the development of an adequate degree of venous outflow resistance [11,12]. A full erection is accompanied by an absence of in- and outflow of blood in the corpora cavernosa [11]. Therefore, measurements of penile blood flow have gained interest as an alternative methodology for non-invasive erection detection. European Association of Urology guidelines state that patients with low flow priapism have blood saturation drops in the corpora cavernosa up to values below 30 (mmHg) during this pathological erection [13,14]. Blood saturation decreases are also expected to occur during normal erection. However, no studies have investigated this hypothesis.

A non-commercial penile sensor was developed to measure hemoglobin concentration of penile tissue. This sensor measures differences in the light reflection of hemoglobin (LRH) of the penis. It was chosen to first investigate the ability of detecting LRH values before a SpO2 penile sensor will be developed.

This study aims to determine the feasibility of erection detection through measurement of differences in light reflection of hemoglobin (LRH) as a precursor to application of SpO2 measurements to detect erections.

## 2. Materials and Methods

This was an observational, cross-sectional pilot study in patients undergoing penile duplex measurements. The study was reviewed, registered and approved by the ethical committee of the Medical Research Ethics Committees United Nieuwegein, the Netherlands and the boards of all participating centers and it conforms to the provisions of the Declaration of Helsinki. (Registration numbers: NL70945.100.19, R19.074 approved on 17-2-2020). The study was registered on ClinicalTrials.gov number: NCT 06402097.

### 2.1 Participants

Patients, aged 18 - 60 years, with an indication for a duplex of the penis in the outpatient Urology clinic of the Sint Antonius Hospital Nieuwegein, the Netherlands, were eligible for participation. All participants received verbal and written explanations of the study procedures. All participants gave their informed consent for inclusion before they participated in the study. With a questionnaire; demographic data, including age, partner status, comorbidities, surgeries, medications and endothelial risk factors (diabetes, smoking, hypertension, hyperlipidemia) were recorded. An IEFF-5 (International Index of Erectile Function) questionnaire was taken. Patients with sickle cell anemia or patients who were unwilling to sign the informed consent were excluded.

### 2.2 Number of patients

The sample size in this proof-of-concept study was derived from the number of patients that were accepted to undergo a duplex of the penis in our clinic, which were around 40 patients a year. Unfortunately there is no existing literature regarding penile LRH, therefore it is not possible to calculate a sample size. For this pilot test, a sample size of ten patients with a full erection was required to measure differences in LRH. 46,7% of patients have a full erection during duplex measurements [15]. Therefore, a total of 20 patients were included in the study.

### 2.3 Sensing system

The aim of this study was to investigate the differences in LRH values occurring during the measurements of penile erections. The LRH values were measured through a small portable device intended for measuring tissue oxygenation of the penis in a flaccid and erected state. The prototype developed sensor version 0.1 uses a setup based on a development platform for measuring vital signs. The sensor’s hardware is based on existing technology that is used in for example smart watches. The sensor is converted for this study to a sensing system that can be applied in penile measurements.

The device is designed to investigate hemoglobin levels in the penis through realtime monitoring with two receiving photodiodes and one emitting LED (light emitting diode) with a wavelength of 536 nanometer coupled to a microprocessor. The two optical photodiodes measure reflected light and ambient light from the penis. The illuminating LED makes it possible to measure a difference in the light reflection of hemoglobin. Although the transversion of light-transmission values to SpO2 requires a second LED, it was chosen to first investigate the ability of detecting LRH values before a SpO2 sensor for the penis will be developed. See Figure 1 for an impression of the device.

**Figure 1.**
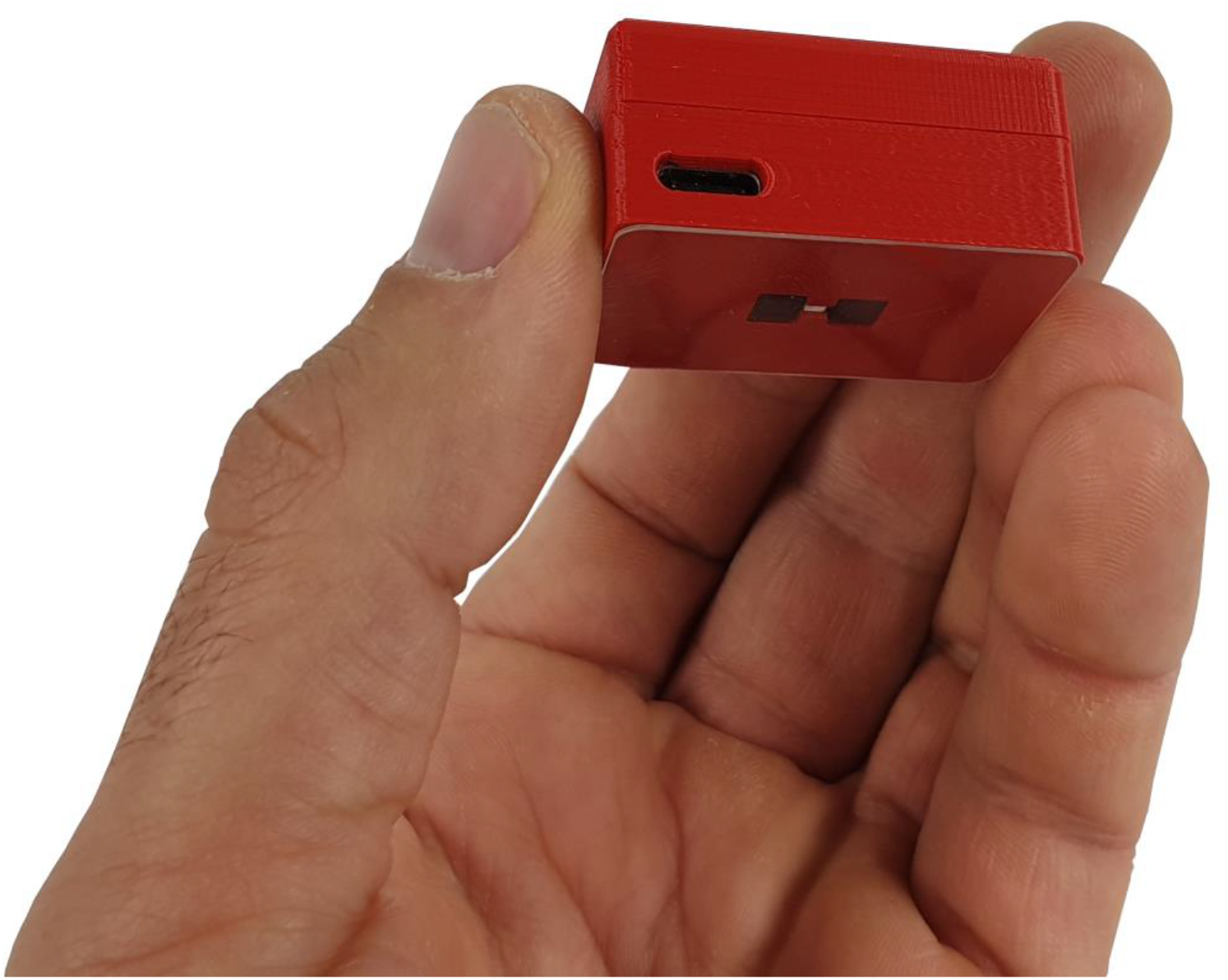
Picture of the penile self-developed sensor (measuring 41 × 27 x 19 mm)

The sensor settings are based on the information from the sensor description of the functionality options, set on measuring underneath the skin of the penile shaft [16]. The settings are configured as follows: both photodiodes enabled, utilizing ambient light cancellation, with a pulse width of 58 microseconds, a sample rate of 100 per second, operating in dual pulse mode. The MAX86141 Evaluation System (Maxim Integrated, Analog devices, San Jose, California, United States) [16] is the heart of the sensor and is incorporated in a body (measuring 41 × 27 x 19 mm) made of the antibacterial nanocomposite PlactiveTM (Copper3D, Omaha, Nebraska, United States) [17]. The system also contains a Bluetooth Low Energy unit and a Lithium-ion polymer rechargeable battery. The sensor monitors a clock, calendar and data storage. The device is manufactured by the researcher in the Netherlands, Utrecht.

### 2.4 Duplex

Duplex ultrasound of the penis is done to obtain ultrasound images and vascular data of the corpora cavernosa (Hitachi Aloka V60 penile probe L55 13-5, Hitachi, Tokyo, Japan). Interpretation of the data is done by a urologist and a nurse who are specialized in ultrasounds and duplex examinations of the penis. The resistance index (RI), peak systolic velocity (PSV) and a clinical conclusion are noted by the examiner.

### 2.5 Study procedure

A schematic flowchart of the study design is shown in Figure 2. First, a duplex ultrasound of the penis was done. After the first assessment the LRH sensor was placed on the left dorsal side of the penis (Figure 3). After intracavernous injection of prostaglandin E1 (20 uG alprostadil) the duplex ultrasound assessment was completed. After the occurrence of full rigidity, the LRH sensor was placed on the same side on the penis to identify LRH values during erection. All measurements were performed by the principal investigator, who pressed the sensor against the penile shaft. The patients without full rigidity after injection with prostaglandin E1 were excluded and measurements with the LRH sensor were aborted. Duplex measurements were still conducted for diagnostic purposes.

**Figure 2.**
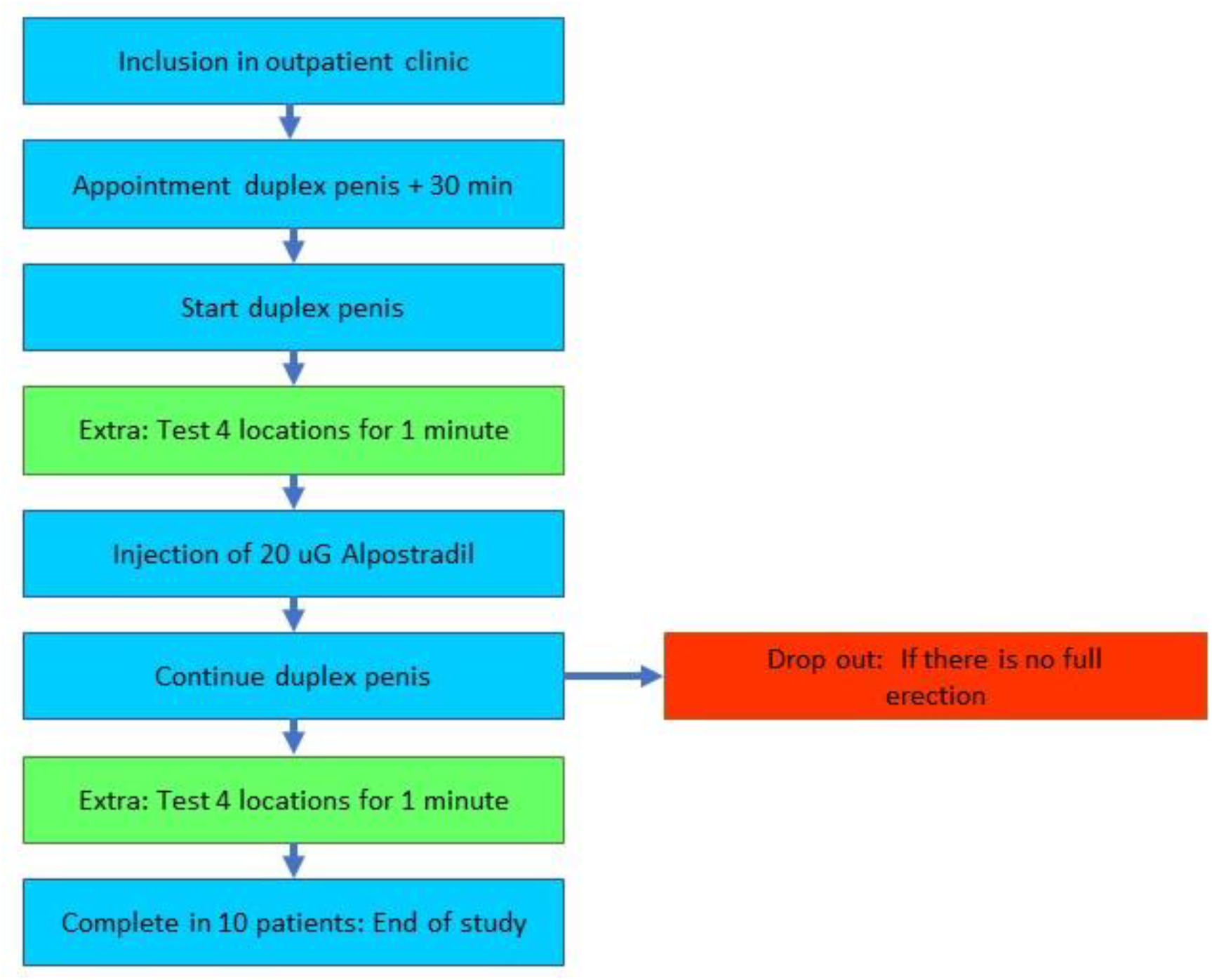
A schematic flowchart of the study design

**Figure 3.**
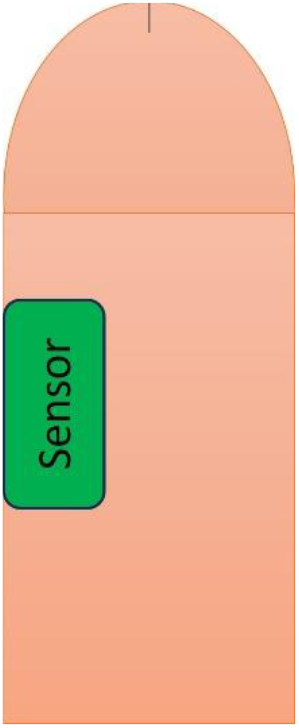
A schematic representation of how the sensor is placed on the penis; top view of the penis with the sensor

### 2.6 Data analysis

Raw data is send from the microprocessor to a computer via a Bluetooth (low energy) connection for further configuration, real-time monitoring, data processing and storage. The data is processed to Microsoft Excel version 2211. The mean LRH value for the measured location before and during an erection was determined. Per second there were 5,25 measurements, for each measured location the average value was taken from the 80 LRH signals, this is around 15 seconds of the measurement. Outcomes were analyzed in IBM SPSS statistics 25 (SPSS inc. Chicago, IL, USA). Comparison of the LRH values before and during erection was tested for significant differences with a paired t-test. Differences were considered significant for alpha <0.05 for the entire study.

For each patient demographic data including age, BMI, partner status, comorbidities, surgeries, medications and endothelial risk factors (diabetes, smoking, hypertension, hyperlipidemia) were recorded. Furthermore, the IIEF-5 (International Index of Erectile Function) questionnaire was conducted (See Appendix A) [18]. Descriptive statistics were given with a mean (SD) or the number of patients and percentages for these data.

## 3. Results

Between June 5th 2020 and September 25th 2020, twenty patients were eligible for participation in this study and gave an informed consent for inclusion. A total of ten patients achieved full rigidity during prostaglandin E1 induced erection, only the outcomes of these ten patients are considered. An overview of the patient characteristics are shown in Table 1. The average age for these patients is 45.4 years (SD 11). The average BMI is 25.6 kg/m^2 (SD 1.7). Of the included patients, 30% suffered from endothelial risk factors, this was noted in the patients records. The IIEF-5 score ranges from 2 to 22 with a mean average of 13 (SD 6.1).

**Table 1.** Baseline patient characteristics (n=10).

| Characteristics | Patients with full rigidity |
| --- | --- |
| Age (years) | 45.4 (11.0) |
| BMI (kg/m<sup>2</sup>) | 25.6 (1.7) |
| Endothelial risk factors n (%) | 3 (30) |
| Diabetes | 0 |
| Smoking | 3 (30) |
| Hypertension | 0 |
| Hyperlipidemia | 2 (20) |
| Use of medication (not specified) n (%) | 3 (30) |
| Operations (not specified) n (%) | 2 (20) |
| IIEF-5 (combined) (score) | 13 (6.1) |
| Question 1 | 2.4 (0.9) |
| Question 2 | 2.7 (1.3) |
| Question 3 | 2.5 (1.4) |
| Question 4 | 2.9 (1.4) |
| Question 5 | 2.5 (1.6) |
| Partner status n (%) |  |
| Yes | 7 (70) |
| No | 3 (30) |
| Duplex |  |
| RI | 0.91 (0.1) |
| PSV (cm/s) | 30.9 (6.8) |
| Conclusion n (%) |  |
| Arterial | 4 (40) |
| Venous | 1 (10) |
| No abnormalities | 5 (50) |

\* Values are means ± SD, or number and (%) of participants. BMI, body mass index; IIEF-5, International Index of Erectile Function[18]; RI, resistance index; PSV, peak systolic velocity.

The outcome of the duplex measurement show a maximal RI in patients ranging from 0.7 and 1 with an average of 0.9. PSV is on average 31 cm/s (SD 6.8). Diagnostic outcomes of the duplex measurements were an arterial cause for ED in four patients, a venous cause in one patients and in five patients no abnormalities were found.

The outcomes of the LRH values during the flaccid state were on average 288,994 (97,874) and during erection 322,515 (119,260). The difference was 33,521. The paired t-test shows no significant differences in penile LRH values between flaccid and erect state (p = 0.11).

## 4. Discussion

This study presents the results of the first investigation to evaluate the non-invasive measurements of hemoglobin light reflection with a penile LRH sensor, a precursor of a SpO2 penile measurement. An increase in LRH was found during an erection. Despite a lack of significant effect, a non-invasive LRH sensor does have the potential to contribute to erection detection. It is expected that a new penile SpO2 sensor could assist the non-invasive erection detection after further research.

Determining blood gas saturation values during this study would have given further insight into the validity of the results of this study. However, the invasiveness of such investigation does not outweigh the clinical implications of the outcomes. Brown et al. [19] found a lower cavernosal oxygen tension in men with arteriogenic impotence evaluated with invasive penile cavernosal blood gas levels with a flaccid penis. Padmanabhan et al. [20] measured penile blood saturation in a flaccid penis after the application of medication to induce an erection. This study described an increase in saturation 10-15 minutes after injection [20]. However, it was not mentioned what the strength and duration of the erections were in these patients during the measurement. Although several studies have been conducted on penile saturation in a flaccid state, this is the first study measuring LRH in ED patients with a non-invasive sensor during an erection [19,20]. This is the first step to improve the non-invasive measurements for patients with ED and a precursor for the development of a penile SpO2-sensor.

It was noticed that, the erection weakened in some patients during the measurement, which was not expected. This weakening could have decreased the measured change in LHR values between flaccid and erectile state. In normal physiology the ending of penile rigidity is preceded by the inflow of fresh blood in the corpora cavernosa. At that moment causing a change in hemoglobin concentration and thus a change in LRH values was expected. For future studies we propose to test the sensor during nocturnal erections in healthy volunteers, to tackle the problem of ambiguity about the state of the erection by combining it with simultaneous data from a Rigiscan measurement.

This study showed that 50% of patients had an erection induced by prostaglandin E1 injection, which was comparable with 47 % found by Iribarren et al [15]. The premature loss of erection could be explained by the patient selection for duplex, which was intended for patients with presumed organic erection failure. Various factors could have further caused the erection loss; the presence of an extra sensor, doing research and an extra examiner in the room causing more sympathetic activation which caused higher adrenaline levels. This can prevent the veno-occlusive function of the corpora cavernosa and give less response to the vasoactive agent [20,21].

Slight differences between the LRH values could be explained by the pressure effect of sensor placement. Furthermore, differences in light reflecting could also be caused by deviations from penile hair and penile curvature. If, for one of those reasons, the sensor is slightly different placed, this can lead to different values. In future research, the sensor application must be standardized in one location to ensure the reproducibility of outcomes. A LRH penile sensor is a first step in developing a device that can measure cavernous tissue oxygenation with a pulse oximeter. Future studies should focus on adding an extra LED to enable conversion of the data into the clinically used percentile saturation values. Furthermore, the results of this study could be validated by overnight measurements in healthy volunteers, because these have several good erections a night [22]. It is crucial that the placement of the sensor is standardized for accurate outcome interpretation. These overnight measurements would be the first step in determining the value of penile blood saturation measurements in the differentiation between a erected and flaccid penis. This pilot study provides an opportunity for further research to identify an objective, minimally invasive evaluation of penile hemodynamics.

## 5. Conclusions

This is the first human pilot study to demonstrate that a higher LRH during the onset of an artificially induced erection in patients with ED can possibly be detected using LRH. The study demonstrates a LRH penile sensor to be a potential novel and non-invasive method of measuring erections. Further research is needed with the penile LRH sensor before the development of a SpO2 sensor and to test the feasibility of the application of the sensor for nocturnal erection detection. The study is a promising advancement towards the further development of a much-needed non-invasive sensor that has the potential to revolutionize the diagnosis of ED.

## Author Contributions

EJT: conceptualization, methodology, formal analysis, investigation, data curation, writing and project administration. HJT: reviewing and editing. HWE, RCMP and JJHB: conceptualization, supervision, reviewing and editing. All authors have read and agreed to the published version of the manuscript.

## Institutional Review Board Statement

The study was conducted in accordance with the Declaration of Helsinki, and approved by the Dutch Medical Ethics Committee was consulted and an application was submitted, ethical approval was given for this study (NL70945.100.19, R19.074 approved on 17-2-2020). The study was registered on ClinicalTrials.gov number: NCT 06402097.

## Informed Consent Statement

Informed consent was obtained from all subjects involved in the study.

## Data Availability Statement

The data presented in this study are available on request from the corresponding author. The data are not publicly available due to privacy.

## Acknowledgments

Not applicable.

## Appendix A

The International Index of Erectile Function (IIEF-5) Questionnaire [18] Over the past 6 months: The IIEF-% score is the sum of questions 1 to 5. The lowest score is 5 and the highest score is 25. Lower scores indicate higher perceived erectile dysfunction and higher values indicate lower perceived erectile dysfunction.

